# Longevity of SARS-CoV-2 immune responses in haemodialysis patients and protection against reinfection

**DOI:** 10.1101/2021.01.22.21249865

**Authors:** Candice L Clarke, Maria Prendecki, Amrita Dhutia, Claire Edwards, Virginia Prout, Liz Lightstone, Eleanor Parker, Federica Marchesin, Megan Griffith, Rawya Charif, Graham Pickard, Alison Cox, Myra McClure, Richard Tedder, Paul Randell, Louise Greathead, Mary Guckian, Stephen P. McAdoo, Peter Kelleher, Michelle Willicombe

## Abstract

**Background:** Patients with end stage kidney disease (ESKD) receiving in-centre haemodialysis (ICHD) have had high rates of SARS-CoV-2 infection. Following infection, ICHD patients frequently develop serological evidence of infection, even with asymptomatic disease. The aim of this study is to investigate the durability and functionality of immune responses to SARS-CoV-2 infection in ICHD patients.

**Methods:** Three hundred and fifty-six ICHD patients were longitudinally screened for SARS-CoV-2 antibodies and underwent routine PCR-testing for symptomatic and asymptomatic infection. Patients were screened for nucleocapsid protein (anti-NP) and receptor binding domain (anti-RBD) antibodies. Patients who became seronegative at 6 months were investigated for SARS-CoV-2 specific T-cell responses.

**Results:** One hundred and twenty-nine (36.2%) patients had detectable antibody to anti-NP at Time 0, of which 127 (98.4%) also had detectable anti-RBD. At 6 months, of 111 patients tested, 71(64.0%) and 97 (87.4%) remained anti-NP and anti-RBD seropositive respectively, p<0.001. For patients who retained antibody, both anti-NP and anti-RBD levels reduced significantly after 6 months. Ten patients who were anti-NP and anti-RBD seropositive at Time 0, had no detectable antibody at 6 months; of which 8 were found to have SARS-CoV-2 antigen specific T cell responses.

Independent of antibody status at 6 months, patients with baseline positive SARS-CoV-2 serology were significantly less likely to have PCR confirmed infection over the following 6 months.

**Conclusions:** ICHD patients mount durable immune responses 6 months post SARS-CoV-2 infection, with <3% of patients showing no evidence of humoral or cellular immunity. These immune responses are associated with a reduced risk of subsequent reinfection.

**SIGNIFICANCE STATEMENT:** Following infection with SARS-CoV-2, patients with end stage kidney disease (ESKD) frequently develop serological evidence of infection, even with asymptomatic disease. Patients with ESKD receiving in-centre haemodialysis (ICHD) have had high rates of SARS-CoV-2 infection. What is not known is how durable the serological responses in ESKD patients are or whether evidence of prior immune responses protect patients from reinfection. In this study of 356 ICHD patients, at 6 months following the detection of SARS-CoV-2 antibodies, fewer than 3% of patients lacked evidence of either humoral or cellular immunity. Furthermore, patients with serological evidence of infection had a significantly lower risk of being diagnosed with subsequent infection or ‘reinfection’, suggesting functional immune protection.

## Introduction

Efficacy results from several SARS-CoV-2 vaccine trials was welcome news at the end of 2020, as the roll out of effective vaccination programmes set to mark the beginning of the end of the pandemic(1-3). The Moderna (mRNA-1273), Pfizer/BioNTech (BNT162b2 mRNA) and Oxford/AstraZeneca (ChAdOx1 nCoV-19) vaccines have all been shown to induce robust humoral and cellular immune responses against the spike protein of the SARS-CoV-2 virus, which importantly protect individuals from risk of subsequent infection(4, 5). However, given the logistical issues associated with supply, distribution and administration of vaccines globally; adjunct prevention and control measures are going to need to be continued in the months to come.

Patients with end stage kidney disease (ESKD) have been identified as having a poor prognosis following SARS-CoV-2 infection (6-8). In addition, it is also recognised that patients receiving in-centre haemodialysis (ICHD) are at higher risk of acquiring infection due to the inability to shield effectively(8). Using serological methods, we have previously shown that ESKD patients readily seroconvert following confirmed SAR-CoV-2 infection; we have also shown that asymptomatic seroconversion is common in the high exposure setting of ICHD units (7). What is not currently known in this population, is the durability of detectable immune responses and whether the presence of SARS-CoV-2 antibodies protects an individual with ESKD from reinfection.

In this study, we report the longitudinal serological status of a large cohort of ICHD patients. The aim of our study was to compare the longevity of the antibodies to the different SARS-CoV-2 antigenic targets, namely the nucleocapsid and receptor binding domain of the spike protein. We investigate cellular immune responses in patients in whom antibody responses have waned, and finally we evaluate whether immune responses to SARS-CoV-2 infection protect dialysis patients from subsequent reinfection.

## METHODS

### Patient Selection

Three-hundred and fifty-six patients receiving ICHD within 2 units affiliated with Imperial College Renal and Transplant Centre as previously reported, were included(7). Patients were followed up from 24^th^ February 2020 until 1^st^ January 2021. All patient samples (n=356) at time 0 were tested for N-protein and RBD antibodies. At 6 months, all patient samples (n=301) were tested for N-protein antibodies; samples which were either anti-NP+ or indeterminant were tested for RBD antibodies. Patient outcomes, including all new SARS-CoV-2 infections, confirmed by viral detection, were recorded up until the 1^st^ January; which incorporates data from the 2^nd^ wave of infections in the UK.

The study was approved by the Health Research Authority, Research Ethics Committee (Reference: 20/WA/0123 - The Impact of COVID-19 on Patients with Renal disease and Immunosuppressed Patients).

### SARS-CoV-2 antibody detection

Baseline serum from all patients were tested for Nucleocapsid protein (NP) using the Abbott Architect SARS-CoV-2 IgG 2 step chemiluminescent immunoassay (CMIA) assay according to manufacturer’s instructions. For this study, samples were interpreted as positive or negative according to the manufacturer’s instructions with a cut off index value of 1.4(9). All baseline serum underwent testing for total receptor binding domain (RBD) antibodies using an in-house double binding antigen ELISA (Imperial Hybrid DABA; Imperial College London, London, UK), which detects total RBD antibodies(10). The in house assay cut off was calculated from receiver operating characteristic curve analysis, and serum reactivity normalized by using the signal-to-cutoff ratio (S/CO); the ratio of optical density values generated in a sample to the cutoff optical density value. For this study, a sample was considered antibody positive if the S/CO was >1.2. At 6 months all samples were re-tested for NP antibodies. Samples with positive or equivocal NP antibodies (cut off-index (0.25-2.5)) were tested for RBD antibodies.

### Detection of SARS-CoV-2 T-cell responses

SARS-CoV-2 specific T-cell responses were detected using the T-SPOT^®^ *Discovery* SARS-CoV-2 (Oxford Immunotec) according to the manufacturer’s instructions. In brief peripheral blood mononuclear cells (PBMCs) were isolated from whole blood samples using the *T-Cell Select*™(Oxford Immunotec) where indicated. Then 250,000 PBMCs were plated into individual well of a T-SPOT^®^ *Discovery* SARS-CoV-2 plate. The assay measures immune responses to 5 different overlapping SARS-CoV-2 structural peptide pools; S protein (spike protein), N protein (nucleocapsid protein), M protein (membrane), S1 and S2 spike protein components as well as positive and negative controls. Cells were incubated and interferon-γ secreting T cells were detected. The sum of T-SPOT immune responses to SARS-CoV-2 structural peptides was calculated. Counts more than 12 spots per 250,000 PBMC were reported as positive(11).

### Diagnosis of SARS-CoV-2 infection

Infection with SARS-CoV-2 was confirmed through reverse-transcriptase polymerase chain reaction (RT-PCR) assay of nasopharyngeal swab specimens, either following routine screening or acute presentation. Reverse-transcriptase PCR was carried out as per Public Health England (PHE) guidelines, utilising certification marked assays with primers directed against multiple targets of SARS-CoV-2 genes (12). Routine screening of patients for the development of symptoms or a fever occurred prior to each haemodialysis session from the 9^th^ March onwards. In addition, routine asymptomatic swabbing was performed in June, and recommenced weekly during the second surge from 15^th^ November.

### Statistical Analysis

Statistical and graphical analyses were performed with MedCalc® v19.2.1. The two-sided level of significance was set at p<0.05. Chi-squared testing were used for proportional assessments. Non-parametric data were compared with the Mann-Whitney test. The Wilcoxon test was used to compare antibody levels of paired samples. Using the log rank test, Kaplan-Meier analyses were used to estimate and compare the risk of infection (or re-infection) by serological status. We recorded any positive PCR test at >60 days following a positive serological test at Time 0, to prevent capture of persistent viral detection of the primary infection(13). As we were not routinely PCR swabbing all asymptomatic cases at the time of first serological sampling, we also used only the PCR results taken >60 days post serological screening in the antibody negative group. Subsequent PCR positive free survival was censored for death, in the absence of PCR confirmation and transplantation.

## RESULTS

At Time 0, 129/356 (36.2%) of all patients had detectable anti-NP and 134/356 (37.6%) had detectable anti-RBD. The clinical characteristics of patients by anti-NP status has been described previously and are shown in Table 1(7).

**Table 1.** Patient characteristics by nucleocapsid protein antibody status at Time 0.

| **Variable** |  | **Anti-NP Pos**<br>**N=129 (%)** | **Anti-NP Neg**<br>**N=227 (%)** | **p value** |
| --- | --- | --- | --- | --- |
| **Gender** | **Female** | 47 (36.4) | 83 (36.6) | 0.98 |
|  | **Male** | 82 (63.6) | 144 (63.4) |  |
| **Age** | **Years (median)** | 65 (55-73) | 68 (57-77) | **0.05** |
| **Ethnicity** | **Black** | 18 (14.0) | 28 (12.3) | 0.36 |
|  | **Caucasian** | 29 (22.5) | 61 (26.9) |  |
|  | **Indoasian** | 60 (46.5) | 94 (41.4) |  |
|  | **Other** | 22 (17.1) | 44 (19.4) |  |
| **Cause of ESKD** | **APKD** | 6 (4.7) | 13 (5.7) | 0.90 |
|  | **Diabetic nephropathy** | 48 (37.2) | 86 (37.9) |  |
|  | **Glomerulonephritis** | 19 (14.7) | 42 (18.5) |  |
|  | **Other** | 12 (9.3) | 37 (16.3) |  |
|  | **Unknown** | 38 (29.5) | 42 (18.5) |  |
|  | **Urological** | 6 (4.7) | 7 (3.1) |  |
| **Time at ESKD** | **Years (median)** | 1.7 (0.9-3.2) | 2.2 (0.9-4.0) | 0.18 |
| **Immunosuppressed** | **Yes** | 14 (10.9) | 41 (18.0) | 0.07 |
| **Symptomatic** | **Yes** | 85 (65.9) | 36 (15.9) | **<0.0001** |

### Serostatus and antibody levels at 6 months

Three hundred and one patients had a sample available at 6 months post initial sampling. Of 190 patients who were anti-NP-at Time 0, 6 (3.2%) had detectable anti-NP by month 6; 3 of which had PCR proven disease in the intervening period. In the patients who were anti-NP+ at time 0; the (S/C) was significantly higher in symptomatic patients compared with asymptomatic patients, with a median value of 7.3 (IQR: 6.1-8.5) and 6.2 (3.2-7.1) respectively, p=0.0006. 111/129 patients who were anti-NP+ at Time 0 had a sample available at 6 months. 40/111 (36.0%) were subsequently found to be anti-NP- at 6 months. In patients who had a detectable anti-NP antibody at both time 0 and 6 months; the median (S/C) was significantly lower at 6 months compared with Time 0 at 2.3 (IQR: 0.9-4.3) and 6.9 (IQR: 5.2-8.2) respectively, p<0.0001 (Figure 2).

**Figure 1.**
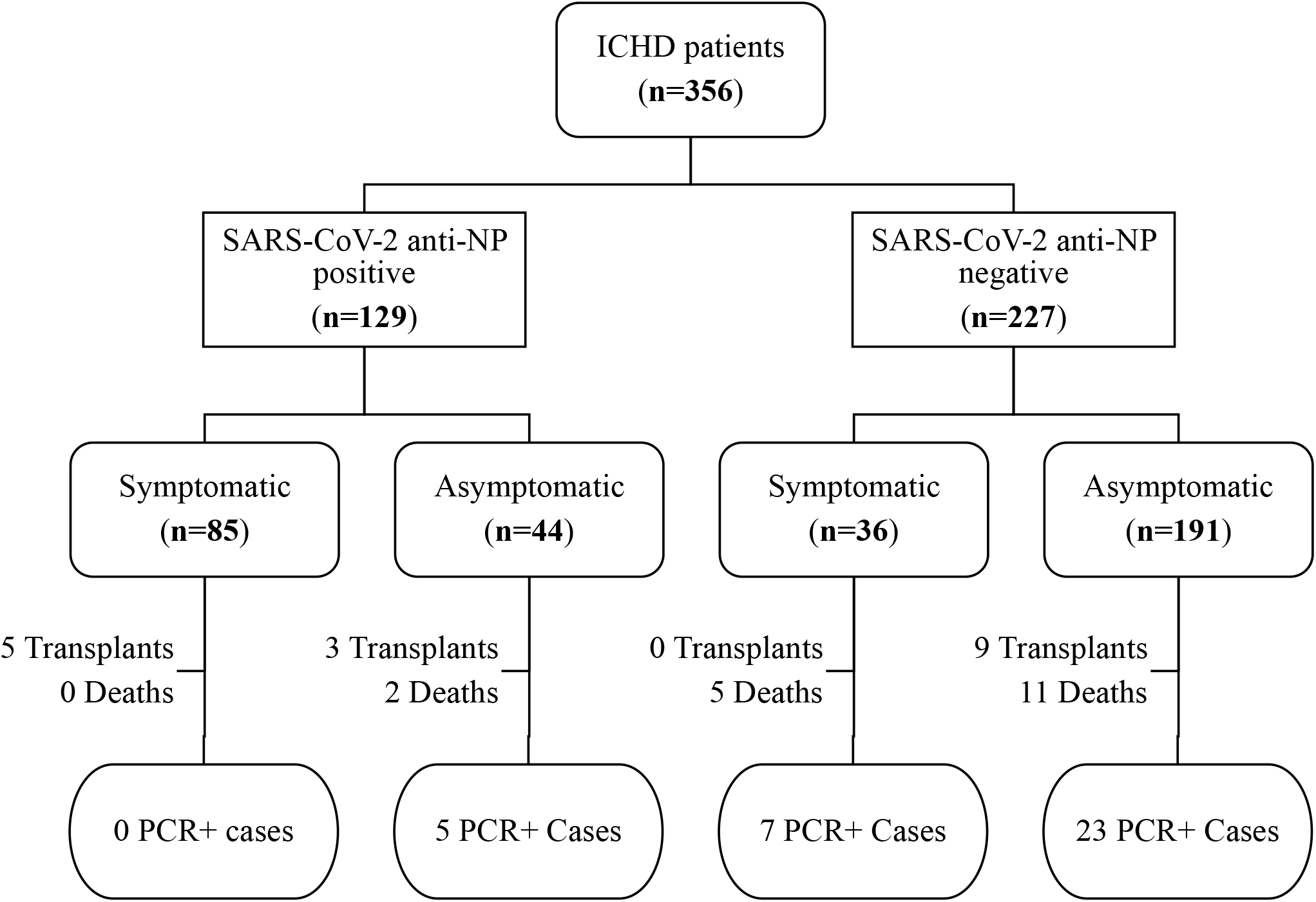
Patient cohort flow diagram by anti-nucleocapsid antibody (NP-Ab) status after first wave of infection.

**Figure 2.**
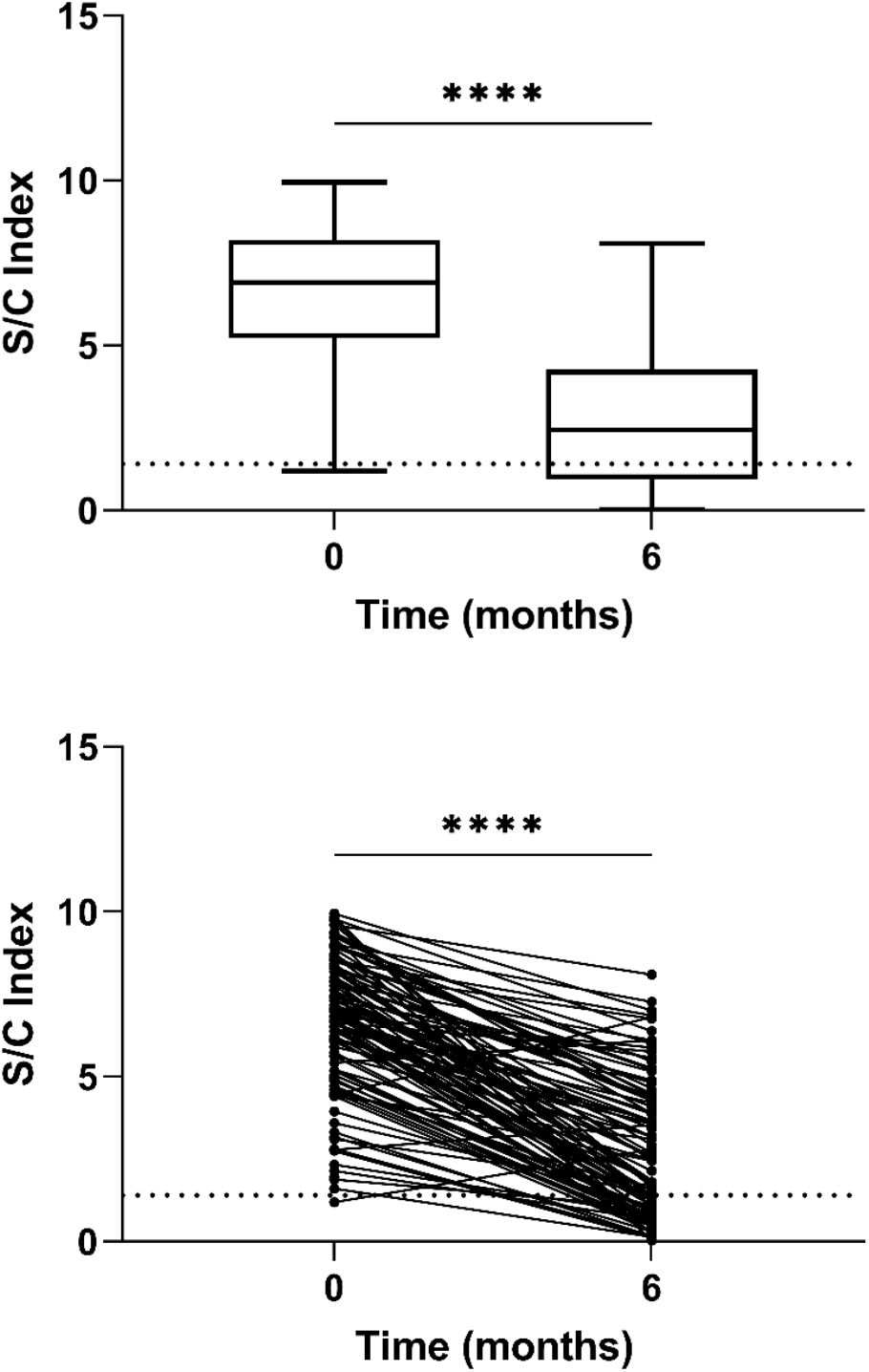
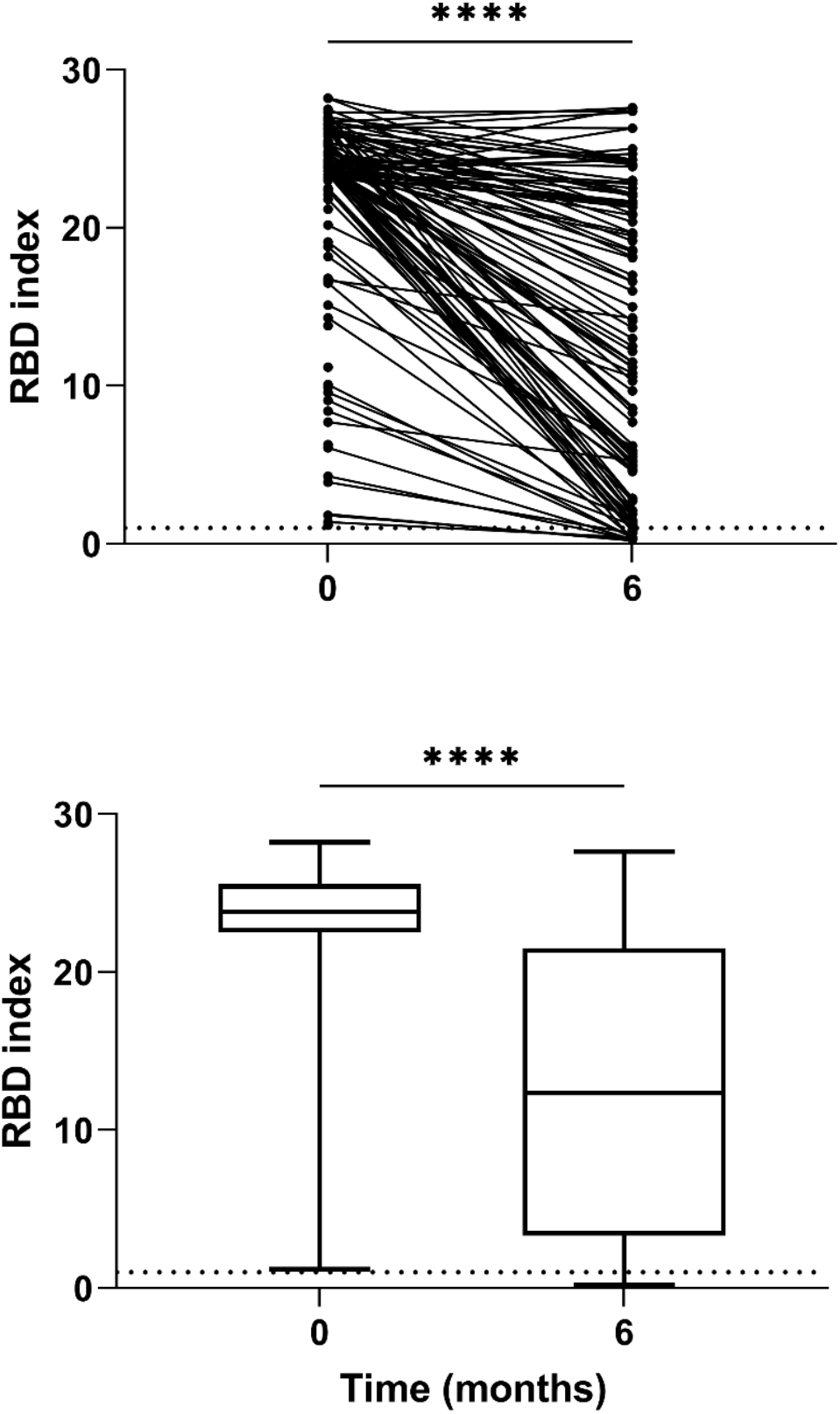
Comparison of Nucleocapsid Protein and Receptor Binding Domain Antibody Levels at Time 0 and Time 6 months. **a. Nucleocapsid Antibody** 40/111 (36.0%) of patients with detectable anti-NP at Time 0 became anti-NP- at 6 months. For those retaining antibodies, the anti-NP index (S/C) was significantly higher at Time 0 compared with 6 months post testing, with a median (S/C) of 6.9 (IQR: 5.2-8.2) and 2.3 (IQR: 0.9-4.3) respectively, p<0.0001. **b. Receptor Binding Domain Antibody** 97/111 (87.4%) of patients with anti-RBD at Time 0 became retained their antibodies at 6 months. Of which, the anti-RBD index (S/C)) was significantly higher at Time 0 compared with 6 months post testing, with a median (S/CO) of 23.9 (IQR: 23.4-26.1) and 23.4 (IQR: 8.9-24.1) respectively, p=0.0004..

**Figure 3.**
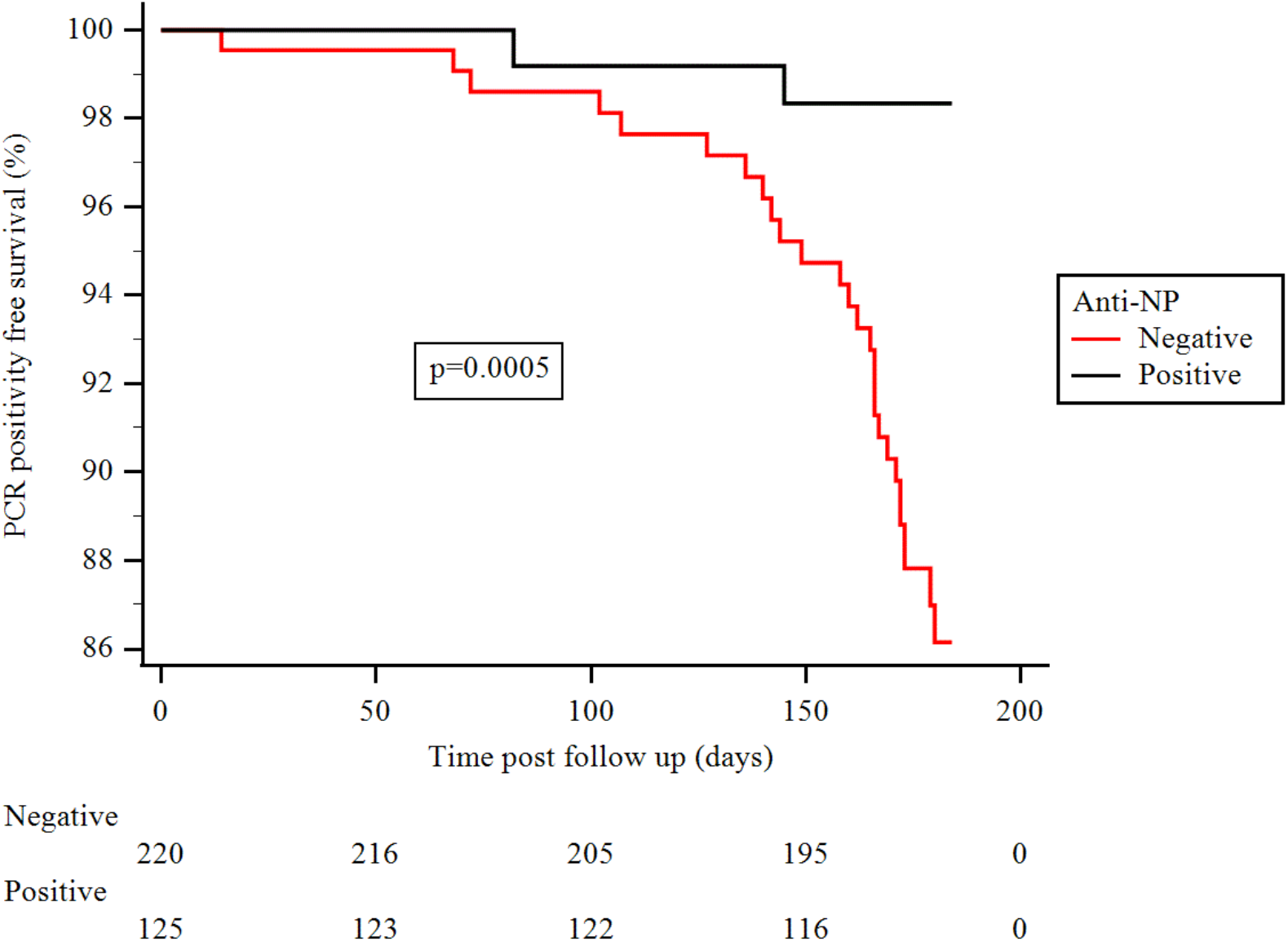
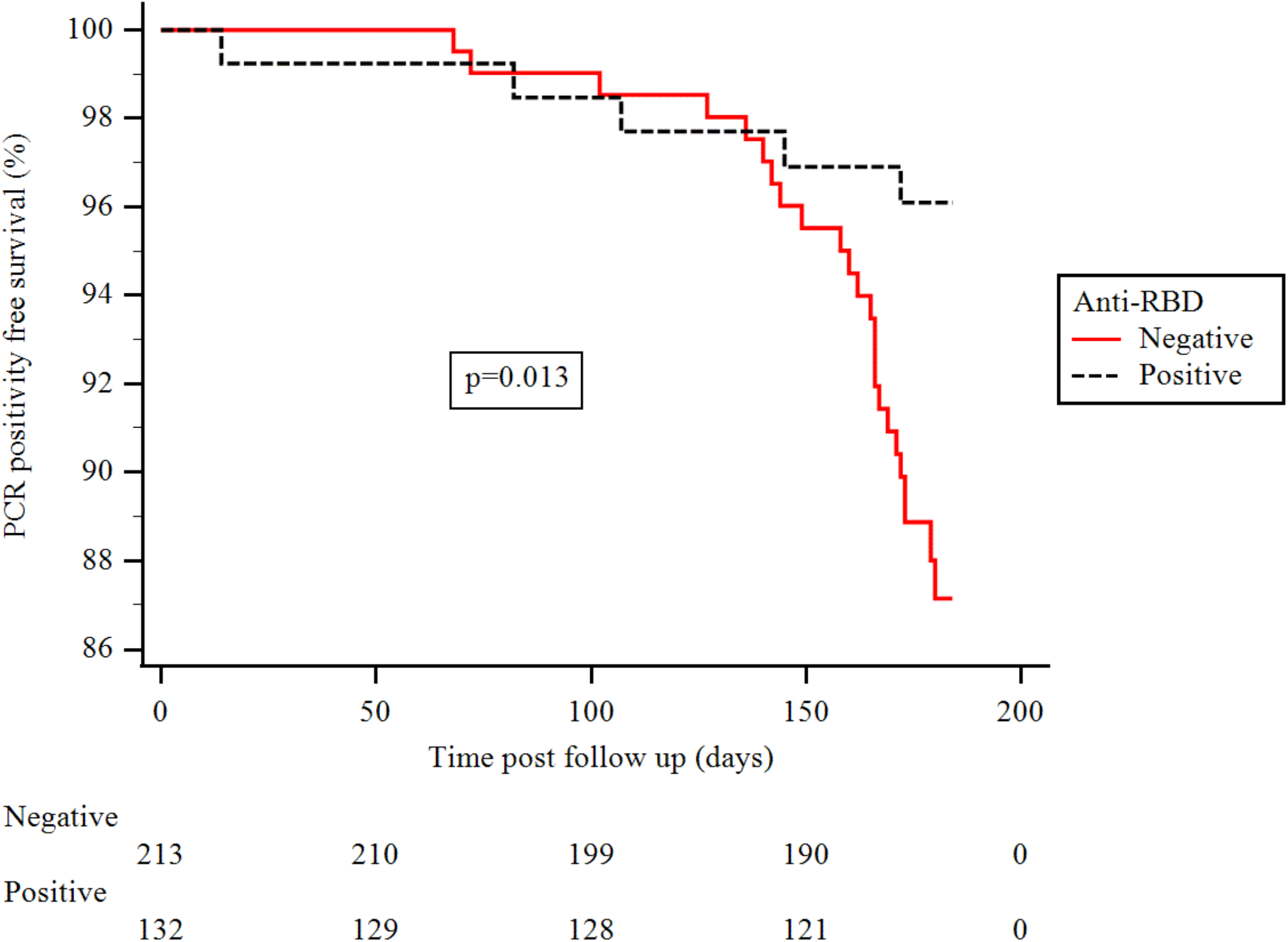
SARS-CoV-2 PCR positive free survival from 60 days following the first serological test by antibody status. **a. Nucleocapsid Antibody** From >60 days after initial serological testing, anti-NP+ patients were at significantly lower risk of being diagnosed with SARS-CoV-2 compared with anti-NP-patients, p=0.0005 (log-rank) **b. Receptor Binding Domain Antibody** From >60 days after initial serological testing, anti-RBD+ patients were also at lower risk of being diagnosed with SARS-CoV-2 by PCR testing compared with anti-RBD-patients, p=0.0051 (log-rank)

Of the 129 patients who were seropositive for anti-NP at Time 0, 127 (98.4%) patients had detectable anti-RBD. Of the 227 patients who were anti-NP- at Time 0, 7/227 (3.1%) had detectable anti-RBD. In anti-RBD+ patients, the antibody index at Time 0 was also significantly higher in symptomatic compared with asymptomatic patients, with a median (S/CO) of 23.9 (IQR: 23.4-26.1) and 23.4 (IQR: 8.9-24.1) respectively, p=0.0004. Of the 111 patient samples available for antibody testing at 6 months, 97 (87.4%) patients remained anti-RBD positive. Anti-RBD durability was significantly superior compared with anti-NP, p<0.0001, and of the 40 patients who became anti-NP seronegative, 28/40 (70.0%) of patients remained anti-RBD+ at 6 months. Similarly to anti-NP, for those patients retaining antibody, the anti-RBD index value was significantly higher at Time 0 compared with 6 months, with a median (S/CO) of 23.8 (IQR: 23.3-25.4) and 12.4 (IQR: 3.8-21.5), p<0.0001.

### T-cell responses at 6 months

Twelve patients who anti-NP+ at Time 0, were seronegative for both anti-NP and anti-RBD at 6 months. T-cell responses were investigated in 11 of these patients. One of these patients who was anti-RBD- at both Time 0 and 6 months, was also found to have unreactive antigen specific T-cell responses. Of the remaining 10 patients, 8 had positive ELISPOT readouts as shown in Table 2. Both patients with a negative ELISPOT test were over the age of 70 years, one had a history of bladder cancer, but neither were iatrogenically immunosuppressed. T-cell responses were not available on patients who were anti-NP-but anti-RBD+ at Time 0.

**Table 2.** Characteristics of 10 patients who were seronegative by nucleocapsid and receptor binding domain antibody at 6 months.

| GENDER | AGE RANGE | ETHNICITY | CAUSE ESKD | 1<sup>st</sup> DIAGNOSIS | 1<sup>st</sup> SEROLOGY |  |  |  | ELISPOT READ OUT | SUBSEQUENT PCR+ |
| --- | --- | --- | --- | --- | --- | --- | --- | --- | --- | --- |
|  |  |  |  |  | Anti-NP | Ab Index | Anti-RBD | Ab Index |  |  |
| M | 40-49 | INDOASIAN | GN | SEROLOGY | + | 5.62 | + | 3.9 | + | NO |
| M | 40-49 | BLACK | HIV | SEROLOGY | + | 1.61 | + | 1.8 | + | NO |
| M | 60-69 | CAUCASIAN | APKD | SEROLOGY | + | 6.16 | + | 16.5 | + | NO |
| M | 60-69 | INDOASIAN | APKD | SEROLOGY | + | 2.76 | + | 1.9 | + | NO |
| F | 60-69 | CAUCASIAN | UNKNOWN | SEROLOGY | + | 2.81 | + | 9.1 | + | NO |
| M | 60-69 | INDOASIAN | DM | SEROLOGY | + | 5.73 | + | 22.4 | + | NO |
| F | 70-79 | OTHER | APKD | SEROLOGY | + | 5.18 | + | 10.1 | - | NO |
| M | 60-69 | INDOASIAN | UNKNOWN | SEROLOGY | + | 7.62 | + | 6.1 | + | NO |
| F | 70-79 | INDOASIAN | DM | PCR | + | 3.32 | + | 4.3 | + | NO |
| M | 70-79 | CAUCASIAN | UROLOGICAL | SEROLOGY | + | 9.21 | + | 23.8 | - | NO |
| M | 70-79 | INDOASIAN | UNKNOWN | SEROLOGY | + | 1.88 | - |  | - |  |

Combining the results of the above immunological assessment, from the original 129 patients who were anti-NP positive, 126 (97.7%) had evidence of persistence of either serological or cellular immune responses against SARS-CoV-2 at 6 months.

### Clinical outcomes associated with seroconversion

Finally, we investigated the clinical relevance of these immune responses, in terms of risk of a subsequent diagnosis of SARS-CoV-2 infection. Within the first 60 days after the Time 0 serological test, 4 anti-NP- and 1 anti-NP+ patients died, and 3 anti-NP- and 3 anti-NP+ patients had a positive PCR test. From >60 days after initial serological testing, anti-NP+ patients were at significantly lower risk of being diagnosed with SARS-CoV-2 compared with anti-NP-patients, p=0.0005 (log-rank), as shown in figure 4. Of the 29 patients who had a positive PCR test at >60 days after the first serological test, 13/29 (44.8%) had follow up for >28 days post PCR testing, of whom 3/13 (23.1%) had died.

Patients with anti-RBD were also at lower risk of being diagnosed with SARS-CoV-2 by PCR testing compared with anti-RBD-patients, p=0.0051 (log-rank), as shown in figure 4. Of the 7 anti-RBD+ patients, who were anti-NP- at Time 0, 2 subsequently went on to have a PCR-positive test at >60 days; the first was diagnosed at day 74 post serological test, and this may represent persistent viral carriage rather than reinfection. The 2^nd^ case was a male in his 70s who was diagnosed at day 112 following a symptomatic infection who has subsequently made a full recovery.

## DISCUSSION

We have shown that immune responses to natural SARS-CoV-2 infection in ICHD patients are durable for up to 6 months, even in patients who had mild or asymptomatic infection. Furthermore, we have provided data that shows that an immune response to SARS-CoV-2 infection may help provide protection against ‘infection’ or reinfection in dialysis patients in the medium term. Using serological status to determine previous exposure in ICHD populations, may therefore help identify patients at higher risk of primary infection whilst awaiting vaccine administration.

A recent large longitudinal study of healthcare workers has shown that the presence of anti-spike or anti-nucleocapsid antibodies was associated with a reduced risk of reinfection over a 6 month period(13, 14). This data is consistent with the relative sparsity of reports of reinfection in the literature(15, 16). However, given this study included healthcare workers, who are likely to be significantly younger and lack co-morbidity, translating these findings to patients with ESKD in the absence of data would be injudicious. However, the findings of the healthcare worker study are supported by results from a separate report which investigated the medium term humoral and cellular responses in SARS-CoV-2 patients with a range of disease severity and co-morbidities, which showed that robust immune responses may persist for at least 8 months post-infection(17). Whilst this may be more comparable data for dialysis patients, a further study from this same research group also showed a disparity in mounting adaptive immune responses in older persons(18). This is of concern for the nephrology community, as dialysis patients are also known to have impairment of both the innate and adaptive immune responses which correlate with premature ageing(19).

Our data, which is the first to investigate longevity of SARS-CoV-2 immune responses in dialysis patients, is therefore of clinical importance. Like others, we have shown that antibody levels wane over time and rate of decay correlates with infection severity(20). However, we acknowledge that use of seroprevalence alone may underestimate ongoing immunity to SARS-CoV-2; with data showing that robust T-cell responses maybe detectable in patients who have had mild or asymptomatic disease, even in the absence of antibodies(21). In our dialysis cohort, we have shown that less than 3% of patients lacked evidence of either serological oor T-cell responses at 6 months post infection. Of upmost clinical relevance, we have also shown that a detectable serological response to SARS-CoV-2 infection, appears to be protective against reinfection in dialysis patients, even at 6 months post-infection. This data is encouraging given the high rates of infection seen during the early stages of pandemic, as whilst dialysis patients await vaccination, previous exposure to infection may offer some protection. It is worth noting that of the 2 patients with serological evidence of anti-NP, who had subsequent viral detection by RT-PCR at days 142 and 205 post initial detection of seroconversion, one patient has HIV and the other patient had previously had a kidney transplant (Supplemental Table 1). In addition, the 2 patients with no serological or cellular evidence of immunity at 6 months, were both over the age of 70 years. Therefore, although we have shown that dialysis patients per se appear to mount a protective immunological response to SARS-CoV-2, there may be additional recognised clinical factors which may impair immunity and influence outcome in dialysis patients, which requires further study.

Whilst we have shown the development and sustainability of post-infection SARS-CoV-2 immune responses, we still do not know whether this high-risk patient population will mount an adequate vaccine response. To date, patients with ESKD, whether on dialysis or transplanted, have been excluded from the SARS-CoV-2 vaccine trials. Dialysis patients are recognised to have lower rates of seroconversion to vaccinations compared with healthy controls, which in part is due to the impact of uraemic toxins on the immune response(22). This also translates into higher sepsis related mortality rates in dialysis patients compared with the general population(23). This immune deficiency appears to be related to responses to new antigenic pathogens, which of vital importance in the outcome to SARS-CoV-2 vaccination(24). As there is no ESKD efficacy data available from any of the SARS-CoV-2 vaccines to date, we can only hypothesise from previous vaccination programmes that seroconversion in dialysis patients will be inferior than the general public. Extrapolating from the knowledge that the immunosenescence associated with aging is seen prematurely in dialysis patients, and that older people had significantly lower levels of anti-spike protein antibody and neutralising antibody levels at 28 days post the BNT162b2 injection than younger patients; suggests the urgent need for some prospective data in this vulnerable population(5, 25). Furthermore, baseline serological status of patients from past exposure may be of relevance in ESKD patients, and it will be of interest to evaluate whether vaccine responses are inferior in infection naïve patients(26).

This study has several limitations, in part due to the time sensitive nature of the results which have led us to take a pragmatic approach to sample processing. The study would have been strengthened by the addition of more laboratory data on viral loads over time. Certainly, there have been case reports of prolonged viral shedding, which may be more common in immunosuppressed patients(27, 28). However, we used a 60-day cut off for ‘new’ PCR positivity, as utilised by others previously(13). It is also relevant to highlight that all patients did subsequently undergo routine asymptomatic PCR swabbing after this time point. A further limitation is that we do not have data available on viral genetic sequencing of infected patients, and it is possible that ‘reinfections’ are due to new variants of SARS-CoV-2, which may evade immune responses to previous strains(29). However, if this was the case, we would have expected to see equal amounts of new variant’ infection’ across all patients. Our serological data may have been strengthened by screening all patients at 6 months for RBD antibodies, which has shown to more closely correlate with neutralising antibody titre(30). Finally, we only performed T-cell ELISPOT assays in selected cases as time and resource limitations prevented further testing at this stage. However, the major strengths of our study, is that to our knowledge this is the first report of longitudinal immunological responses in dialysis patients, which in addition has been correlated with clinical outcome data in asymptomatic and symptomatic infections.

In conclusion, we have shown that dialysis patients mount durable immune responses 6 months post SARS-CoV-2 infection. Less than 3% of infected patients had no detectable serological or T-cell responses at 6 months. We have also shown that risk of subsequent PCR positive confirmed infection was significantly lower in patients who have had detectable antibodies. Together, these data suggest that immune responses post-infection maybe protective against reinfection. Given the high prevalence of primary SARS-CoV-2 infection during the first wave in ICHD units, this is important information for the nephrology community whilst vaccination programmes roll out.

## Data Availability

The authors confirm that the data supporting the findings of this study are available within the article and its supplementary materials.

## ACKNOWLEDGEMENTS

This research is supported by the National Institute for Health Research (NIHR) Biomedical Research Centre based at Imperial College Healthcare NHS Trust and Imperial College London. The authors would like to thank the West London Kidney Patient Association, all the patients and staff at ICHNT (the ICHNT renal COVID group and dialysis staff, and staff within the North West London Pathology laboratories. The authors are also grateful for the support from Hari and Rachna Murgai, and Milan and Rishi Khosla.

## DISCLOSURES

Peter Kelleher and Michelle Willicombe have received support to use the T-SPOT^®^ *Discovery* SARS-CoV-2 by Oxford Immunotec

## Notes

### Competing Interest Statement

The authors have declared no competing interest.

