## Supplemental Table 1 for "Longevity of SARS-CoV-2 immune responses in haemodialysis patients and protection against reinfection"

**Table 1. Characteristics of 5 seropositive patients who were subsequently found to be SARS-CoV-2 positive**

| GENDER | AGE RANGE | ETHNICITY | CAUSE ESKD | IMMUNOSUPPRESSED | 1st DIAGNOSIS | 1st SEROLOGY | | 2nd SEROLOGY | | TIME TO PCR (DAYS) |
|---|---|---|---|---|---|---|---|---|---|---|
| | | | | | | Anti-NP | Anti-RBD | Anti-NP | Anti-RBD | |
| M | 60-69 | INDOASIAN | UNKNOWN | PREVIOUS TRANSPLANT | SEROLOGY | + | + | - | + | 205 |
| F | 70-79 | INDOASIAN | GN | PREVIOUS TRANSPLANT | SEROLOGY | + | + | + | + | 13 |
| F | 70-79 | INDOASIAN | UNKNOWN | HIV | SEROLOGY | + | + | - | + | 40 |
| M | 60-69 | BLACK | GN | HIV | SEROLOGY | + | + | ND | ND | 142 |
| M | 60-69 | INDOASIAN | DIABETES | NO | SEROLOGY | + | + | + | + | 45 |

Shaded columns represent patients who had a positive PCR test <60 days post serology test
